# Lower vaccination coverage against COVID-19 in school-aged children is associated with low socioeconomic status in the Metropolitan Area of Santiago, Chile

**DOI:** 10.1101/2023.03.27.23287800

**Authors:** Enzo Guerrero-Araya, Cesar Ravello, Mario Rosemblatt, Tomas Perez-Acle

**Affiliations:** Fundación Ciencia & Vida, Avenida del Valle Norte 725, Huechuraba, Santiago, Chile; Facultad de Ingeniería, Arquitectura y Diseño, Universidad San Sebastián, Bellavista 7, Recoleta, Santiago, Chile; Facultad de Medicina y Ciencia, Universidad San Sebastián, Lota 2465, Providencia, Santiago, Chile

## Abstract

**Background:** The burden of COVID-19 has been heterogeneous, indicating that the effects of this disease are synergistic with both other non-communicable diseases and socioeconomic status (SES), highlighting its syndemic character. While the appearance of vaccines has moderated the pandemic effects, their coverage has also been heterogeneous, both when comparing different countries, and when comparing different populations within countries. Of note, once again SES appears to be a correlated factor.

**Methods:** To examine the relationship between SES and vaccination coverage, we analyzed publicly available data detailing the percentage of school-aged vaccinated children in different municipalities belonging to the Metropolitan Area (MA) of Santiago, Chile, one of the most largely vaccinated countries in the world. Vaccination data was compiled per school type, either public, state-subsidized and private, at three different dates along the COVID-19 pandemic so to cover the dispersion of *Delta*, and *Omicron*, including *Omicron* subvariants BA.4 and BA.5. We computed the median vaccination ratio for each municipality and school type and calculated their Spearman’s rank correlation coefficient with each one of nine SES indices.

**Findings:** In the MA of Santiago, Chile, the percentage of school-age children who have received vaccinations against COVID-19 correlates with SES. Vulnerable municipalities with low SES exhibit low levels of vaccination coverage. Of note, this strong correlation is observed in both public and state-subsidized schools, but to a meaningless extent in private schools. Although inequity in vaccination coverage decreases over time, it remains higher among students enrolled either in public and state-subsidized schools compared to those of private schools.

**Interpretation:** Available data is insufficient to explore plausible causes behind lower vaccination coverage in vulnerable municipalities in the MA of Santiago, Chile. However, considering the available literature, it is likely that a combination of factors including the lack of proper information about the importance of vaccination, the lack of incentives for children’s vaccination, low trust in the government, and/or limited access to vaccines for lower-income people, may all have contributed to this low vaccination coverage. Importantly, unless corrected, the inequity in vaccination coverage will exacerbate the syndemic nature of COVID-19.

**Funding:** This material is based upon work supported by the U.S. Air Force Office of Scientific Research under award number FA9550-20-1-0196. Financial support is also acknowledged to Centro Ciencia & Vida, FB210008, Financiamiento Basal para Centros Científicos y Tecnológicos de Excelencia de ANID.

## 1 Introduction

As of October 2022, the pandemic produced by the coronavirus associated disease 2019 (COVID-19) has caused more than 6 million deaths worldwide.^1^ After more than two years since the detection of the first case in China,^2^ COVID-19 continues to pose challenges: from the impact of anti-vaxxers movements^3, 4^ to the appearance of new variants such as Delta and Omicron.^5^ In an unprecedented biotechnological response, vaccine manufacturers developed, and Natioinal Regulatory Authorities approved in record time, several vaccines that were proven safe and effective in diminishing infection, hospital admissions, and deaths rates.^6–8^ However, unequal access to vaccines creates enormous risks not only for the population of developing countries but also for the rest of the world. In fact, the Gamma, Delta, and Omicron variants of SARS-CoV2, the etiological agent of COVID-19, emerged from high prevalence conditions in Brazil, India, and the African continent, respectively, spreading rapidly throughout the world.^5^ On top of that, due to the influence of anti-vaxxer movements driven by the abundance of fake news, anti-science, and the promotion of highly individualistic behaviors, the willingness to get vaccinated is a crucial factor to be evaluated and understood so to design better public policies encouraging vaccination.^9–11^

Over the course of two and a half years, COVID-19 has spread across the globe. However, its impact varies considerably depending on the socioeconomic status (SES) of the affected population. Several studies have reported associations between SES and COVID-19 incidence, mortality, and vaccination coverage.^12–14^ Locally, a seminal study showed that during the first stages of the pandemic, mortality attributed to COVID-19 was higher in places with lower SES in the Metropolitan Area (MA) of Santiago, Chile.^12^ This study exhibited data corroborating that COVID-19 is, in fact, a syndemic disease: an epidemiological condition whose burden among the population is synergistic with both non-communicable diseases and SES.^15, 16^ Thus, wealthier municipalities—usually more affluent and therefore healthier—were found to be less vulnerable to COVID-19. A similar correlation has been reported for other countries.^17^ For instance, in the USA, localities with lower levels of education and a higher proportion of African-American population–both factors usually associated with lower SES–are linked with a higher number of COVID-19 cases and fatalities, together with a higher proportion of long-term consequences of the infection.^18–20^ Similarly, in Sweden, a higher number of COVID-19 related deaths, occurred in areas of lower SES,^21^ while in India, population density and literacy have been found to be positively and negatively correlated with COVID-19 infection rates, respectively, highlighting the syndemic nature of this disease.^22^

On top of the association between SES and the number of cases and the poor prognosis of COVID-19, vaccination rates have also been correlated with these indices. Whereas some countries were able to access vaccines from the beginning of 2021 reaching more than 90% of the population, as in the case of Israel and Chile,^23, 24^ others less affluent countries such as Haiti struggle to reach 5% coverage of their population.^23, 25^ On top of that, studies have shown that a lower percentage of the population receiving the COVID-19 vaccine is also associated with lower SES. This association is exacerbated by other factors such as uncontrolled immigration, political instability, and low trust in the government.^9–11, 13, 26^

In addition to the successful vaccination campaign against COVID-19 targeted for the general population,^27^ the Chilean government has rolled out a massive vaccination campaign for children. As of October 2022, more than 90% of the population between 6 and 17 years old was fully vaccinated, having received at least two doses.^28^ In response to governmental approval of COVID-19 vaccines for very young children, a “safe back-to-school plan” was developed by the Ministry of Education, that included lifting seating capacity limitations at schools when a classroom had more than 80% of the students vaccinated.^29^ Notably, even before the completion of the vaccination campaign for children under 12 years old, class attendance restrictions were lifted, eroding the availability of incentives for children to be vaccinated. In contrast, all individuals above 12 years old were compelled by the Ministry of Health’s to have a Sanitary Pass that certified that that person was up-to-date with the vaccination timetable before attending cultural, shopping, and eating venues.^30^

Although the level of vaccination for each school are being closely monitored by the Chilean government, up to our knowledge, the relationship between SES, vaccination coverage and the nature of the school, has not been assessed. In this study, we determined the SES of the 32 municipalities located in the MA of Santiago, the capital of Chile.^31^ For each municipality we computed the median school vaccination coverage per school type, in three dates distributed along the pandemic so to cover the spread of different SARS-CoV-2 variants: November 15, 2021; March 1st, 2022; and May 26, 2022. In doing so, our results indicated a strong correlation between SES indices and the median school vaccination coverage. We further explored differences between school types, evaluating private, subsidized and public schools data separately. As expected, vulnerable municipalities with low SES exhibit lower levels of vaccination coverage. Surprisingly, while a strong correlation between vaccination coverage and SES is present in both public and state-subsidized schools, the correlation is meaningless for private schools. Therefore, in the latter, vaccination coverage seems to be independent of the SES of the municipality.

## 2 Methods

### 2.1 Socioeconomic status

To generate a socioeconomic profile of the municipalities in Santiago, Chile, we used nine different SES indices so to obtain a broader multi-factorial socioeconomic description.

The **Social Priority Index (SPI)**, was published in 2019 by the Regional Ministerial Secretariat of Social Development and Family. It is a synthetic index that integrates relevant aspects of communal social development, including income, education, and health.^32^

The **School Vulnerability Index (SVI)**, published in 2021 by the National Board of School Aid and Scholarships (JUNAEB), is the ratio of the sum of the students in the first, second, and third priority ranking compared to the total school enrollment. Students are classified in priorities 1, 2, and 3 according to poverty conditions and risk of school failure. Thus, students living in lower socioeconomic conditions (priority 1) tend to exhibit a higher risk of school failure, and thus receive the highest priority from the Board.^33^

The **Community Development Index (CDI)**, published in 2020, is a compound index considering three socioeconomic dimensions: **Health and Social Welfare**; **Economy and Resources**; and **Education**, measured by standardized tests. The CDI index, as well as each dimension, is available for each municipality in the country.^34^ Therefore, in this work we decided to consider all indices separately.

The **Poverty, Multidimensional Poverty and People in Households Lacking Basic Services (PHLBS)** indexes are calculated as the percentage of people belonging to each category respectively for each municipality. Poverty is determined by a threshold defined as a function of income level and household size. Multidimensional Poverty is defined as being part of a household that cannot achieve adequate living conditions in a set of five relevant dimensions of well-being: (1) Education; (2) Health; (3) Labor and Social Security; (4) Housing and Environment; and, (5) Networks and Social Cohesion. These conditions are measured through a weighted set of 15 indicators (three for each dimension). Households that accumulate deficiencies of 22*·*5% or more are classified as being in a situation of multidimensional poverty. The Poverty and Multidimensional Poverty indexes were extracted from the National Socioeconomic Characterization Survey (CASEN) 2017, while the PHLBS was extracted from the Social Registry of Households 2020. Both instruments are used by the Ministry of Social Development to focus social assistance among the population.^35^

### 2.2 Vaccinations

Despite in Chile the COVID-19 vaccination campaign is orchestrated and conducted by the Ministry of Health, the Ministry of Education publishes vaccination coverage per grade and school on its website at https://vacunacionescolar.mineduc.cl/. To determine vaccine coverage per school and grade, the Ministry of Education crosses its school enrollment database with that of the vaccination campaign from the Ministry of Health by using the national identity number (RUT) of each student. To study whether the dispersion of different SARS-CoV-2 variants among the population may have incidence on the vaccination process among school-age students in different municipalities in Santiago, Chile, we downloaded data published on November 15, 2021; March 1st, 2022; and May 26, 2022. By selecting these dates we covered the spread of different SARS-CoV-2 variants: *Delta, Omicron* and *Omicron* subvariants BA.4-BA.5, respectively. For each school and grade, data is provided as the percentage of students with full vaccination scheme, incomplete vaccination scheme, and non-vaccinated students per school and grade.

### 2.3 School metadata

The Chilean Ministry of Education maintains publicly available metadata associated with each educational establishment in the following repository https://datosabiertos.mineduc.cl/directorio-de-establecimientos-educacionales/. Metadata was used to identify public, state-subsidized and private schools in each municipality.

### 2.4 Statistical methods

All statistical analyses were performed on the downloaded data by programming in Python v3.1 using the SciPy v1.7.1 package.^36^ To measure inequity, inspired by The Palma ratio,^37^ we used the percentile ratio p90/p40. To do so, we divided the mean vaccination coverage of the 10% (percentile 90) of municipalities with the highest vaccination coverage, by the mean vaccination coverage of the 40% (percentile 40) of municipalities with the lowest vaccination coverage. Importantly, the p90/p40 ratio ensures that maximum equity is achieved when the ratio equals 1. Spearman’s rank-order correlation and multivariate regression were computed using Statsmodels v0.13.1. Plots were made with Seaborn v0.11.2.

### 2.5 Reproducibility

The whole dataset as well as the python coding programmed used to perform the statistcal analyses presented here are available on GitHub in the following repository https://github.com/DLab/SantiagoSchoolVax.

## 3 Results

### 3.1 Exploring the correlation between SES indices: all indices are correlated with SPI

The SPI index has already been used as a proxy for the socioeconomic status of municipalities in Chile.^12^ However, it is only available for the city of Santiago. Therefore, we evaluated the correlation between other SES indices and the SPI to determine whether other SES indices may be used as proxies to access the socioeconomic status of all municipalities in the MA of Santiago, Chile. Our results indicate that all the evaluated SES indices do correlate with SPI (Supplementary Figure 1), with CDI being the one with the highest correlation strength (|*ρ*| *>*= 0*·*90)

### 3.2 Vaccination coverage is correlated with SES in schools belonging to Santiago, Chile

In this study we evaluated the vaccination and SES data of students enrolled in the 1,667 schools belonging to the 34 municipalities of the MA of Santiago, Chile. We divided the population according to the school type: public, state-subsidized and private schools. Their enrollments as of March 2021 were: 297,928 (29%), 575,426 (55%), 169,898 (16%), respectively. The population estimate of Santiago is 6,075,403 (30*·*9% of the country population). As in any other large city around the world, SES indexes vary widely. For instance, the municipality of Providencia (Figure 1A; CDI rank 1) has the lowest rate of Multidimensional Poverty (0*·*034), whilst Lo Espejo (Figure 1A; CDI rank 33), has the highest rate of Multidimensional Poverty (0*·*375): more than 10 times higher than that of Providencia. As expected, the three municipalities with the highest CDI are also the ones with the highest level of vaccination coverage in schools (Figure 1A). In contrast, the four municipalities with the lowest CDI are those with the lowest vaccination coverage, with the exception of Pedro Aguirre Cerda, a municipality exhibiting lower vaccination coverage and ranking 26th in CDI (Figure 1A). Notably, no municipalities with either high or medium vaccination coverage and low CDI could be found. Consequently, no municipalities with low vaccination coverage and high CDI can be found (Figure 1A).

**Figure 1:**
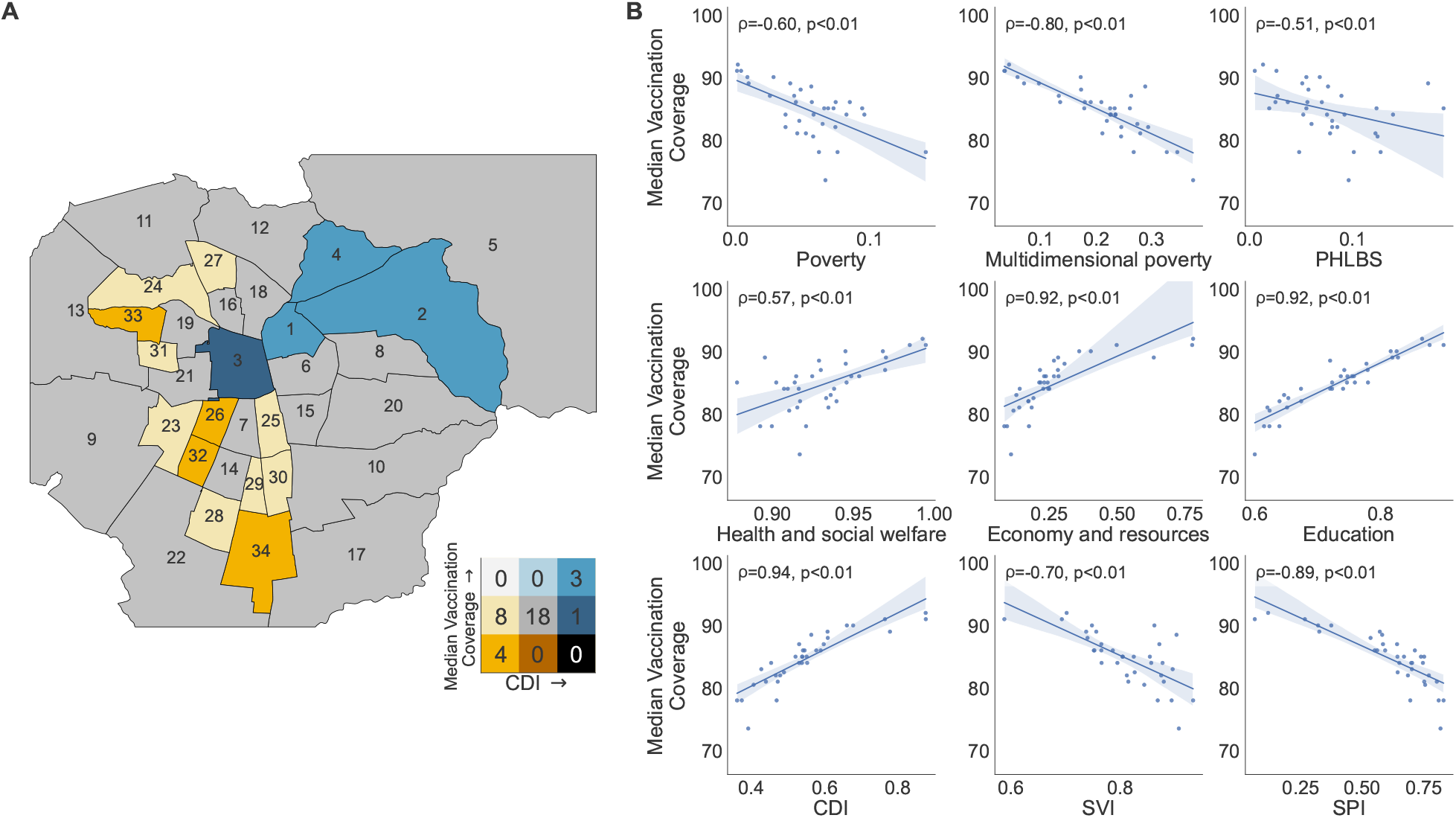
Correlation between school vaccination coverage and SES Indexes in Santiago, Chile as of May 26, 2022. Panel A), municipalities are colored according to their school vaccination coverage and their Community Development Index (CDI) in a Bivariate Map. Municipalities are numbered according to their rank in CDI from higher to lower values. The color coding is shown adjacent to the map using a 3 by 3 matrix where the CDI increases from left to right and the vaccination coverage increases from bottom to top. The values are grouped into three levels. The number labeled in each municipality is sorted by decreasing ranking according to the CDI, as follows: 1, Providencia; 2, Las Condes; 3, Santiago; 4, Vitacura; 5, Lo Barnechea; 6, Ñuñoa; 7, San Miguel; 8, La Reina; 9, Maipú; 10, La Florida; 11, Quilicura; 12, Huechuraba; 13, Pudahuel; 14, La Cisterna; 15, Macul; 16, Independencia; 17, Puente Alto; 18, Recoleta; 19, Quinta Normal; 20, Peñalolén; 21, Estación Central; 22, San Bernardo; 23, Cerrillos; 24, Renca; 25, San Joaquín; 26, Pedro Aguirre Cerda; 27, Conchalí; 28, El Bosque; 29, San Ramón; 30, La Granja; 31, Lo Prado; 32, Lo Espejo; 33, Cerro Navia; 34, La Pintana. (B) The correlation value between six SES Indices and the median school vaccination coverage was evaluated in the 34 municipalities of the Metropolitan Area of Santiago, Chile. *ρ* = Spearman’s rank correlation coefficient; p = p-value.

When analyzing the data gathered on May 26, 2022, three months after the peak of the *Omicron* spreading (around February 14^38^), the correlation between each SES index and the median school vaccination for each municipality ranged from a very strong correlation,^39^ with a |*ρ*| *>*= 0*·*90 for the case of Economy and Resources, Education, CDI, and SPI indices; a strong correlation, with a |*ρ*| *>*= 0.70 for the case of Multidimensional Poverty, and SVI; to a a moderate correlation, with |*ρ*| *>* 0*·*40 for the case of Poverty, PHLBS and Health and Social Welfare. All correlations were statistically significant (*p <* 0*·*01) (Figure 1B). Of note, similar trends with different correlation values between SES indices and vaccination coverage were found on November 15, 2021, three days after the peak of the *Delta* variant spreading,^38^ and in March 5, 2022, in the middle of the second wave of *Omicron* infection produced by the dispersion of the BA.4/BA.5 subvariant^38^(Supplementary Figures 2 and 3).

We further explored whether the correlation between the vulnerability of different municipalities and the vaccination coverage may be accounted for in the three main types of schools existing in Chile: private, state-subsidized and public schools. While parents and tutors enrolling their school-aged children in private schools must pay a full tuition fee, for the case of state-subsidized schools the tuition fee becomes importantly reduced by the state subsidy. On the other hand, for the case of public schools, the state covers the full tuition fee. Our data shows that in both state-subsidized and public schools the same aforementioned correlation can be observed, although correlation values vary (Figure 2A-D). Surprisingly, no correlation between the vulnerability of municipalities and the vaccination coverage for the case of the private school population can be observed (Figure 2E-F).

**Figure 2:**
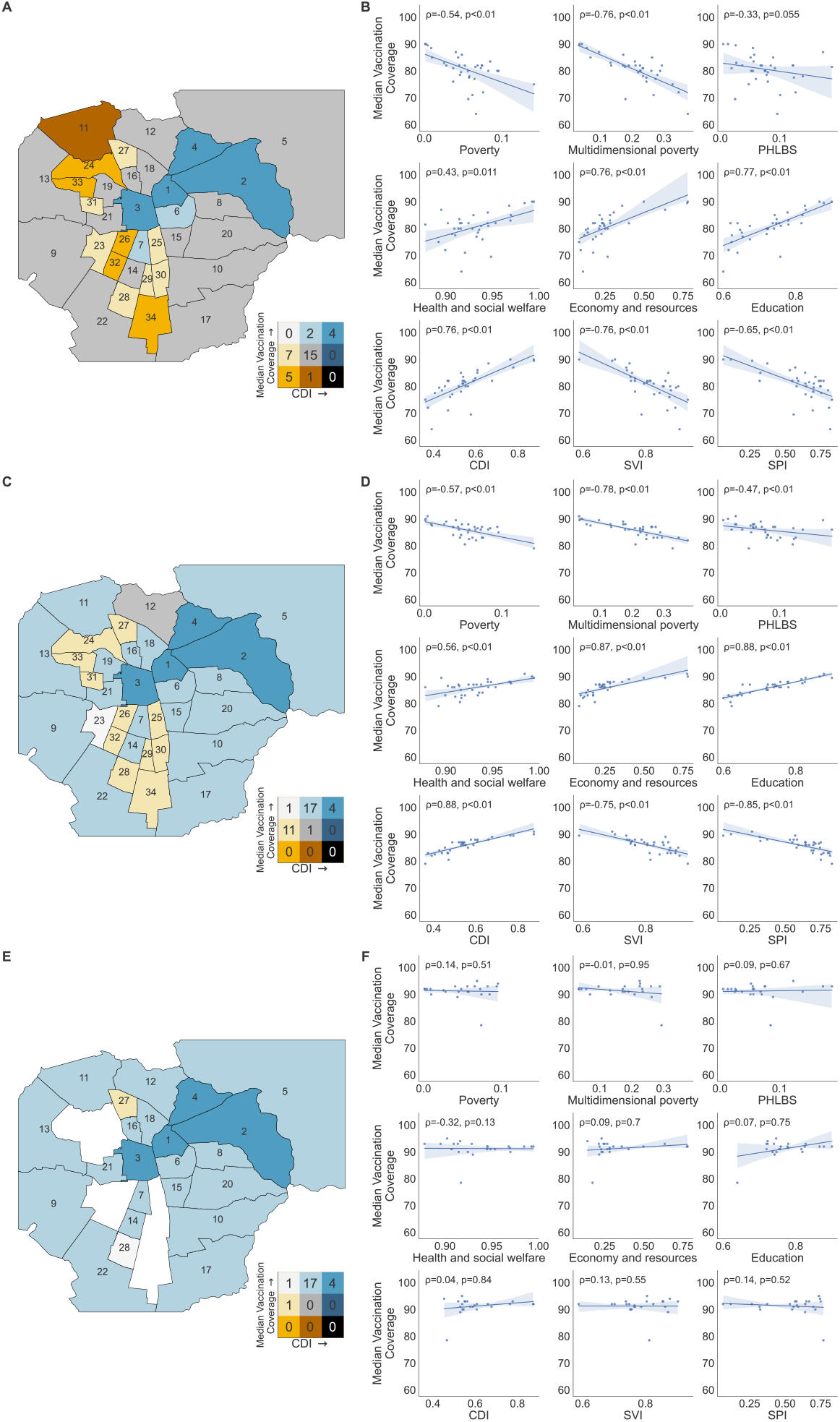
Correlation between school vaccination coverage and SES Indices by school category in the MA of Santiago as of May 26, 2022. All maps represent Municipalities from the MA of Santiago. They appear colored according to their school vaccination coverage and their Community Development Index (CDI) in a Bivariate Map. Municipalities are numbered according to their rank in CDI from higher to to lower values. The color coding is shown adjacent to the map using a 3 by 3 matrix, where the CDI increases from left to right and the vaccination coverage increases from bottom to top. The values are grouped into three levels. The number labeled in each municipality is sorted following a decreasing ranking according to the CDI, as follows. 1, Providencia; 2, Las Condes; 3, Santiago; 4, Vitacura; 5, Lo Barnechea; 6, Ñuñoa; 7, San Miguel; 8, La Reina; 9, Maipú; 10, La Florida; 11, Quilicura; 12, Huechuraba; 13, Pudahuel; 14, La Cisterna; 15, Macul; 16, Independencia; 17, Puente Alto; 18, Recoleta; 19, Quinta Normal; 20, Peñalolén; 21, Estación Central; 22, San Bernardo; 23, Cerrillos; 24, Renca; 25, San Joaquín; 26, Pedro Aguirre Cerda; 27, Conchalí; 28, El Bosque; 29, San Ramón; 30, La Granja; 31, Lo Prado; 32, Lo Espejo; 33, Cerro Navia; 34, La Pintana. Next to each map, the correlation of six SES indices and the median school vaccination coverage is show. *ρ* = Spearman’s rank correlation coefficient; p = p-value. (AB) Public schools data. (CD) Private subsidized schools data. (EF) Private schools data.

### 3.3 Progression of inequity in school vaccination coverage over time

With the aim to assess the inequity of the vaccination coverage, we calculated the p90/p40 ratio in three dates, covering the spread of different SARS-CoV-2 variants. We found that overall, inequity decreases in all schools types over time along the COVID-19 pandemic (Figure 3). Importantly, the largest reduction in the inequity of the COVID-19 vaccination coverage occurs between November 15, 2021 and March 01, 2022, after the peak of the *Omicron* variant dispersion. In all dates evaluated, inequity in public schools remains higher than that of private and state-subsidized schools. Despite a significant correlation between the vulnerability of the municipality and the vaccination coverage does actually exist, denoting that lower SES is correlated with lower vaccination coverage, inequity in vaccination coverage is higher in public schools. On the contrary, both private and state-subsidized schools exhibit higher equity in vaccination coverage, independently of the municipality where the schools are located.

**Figure 3:**
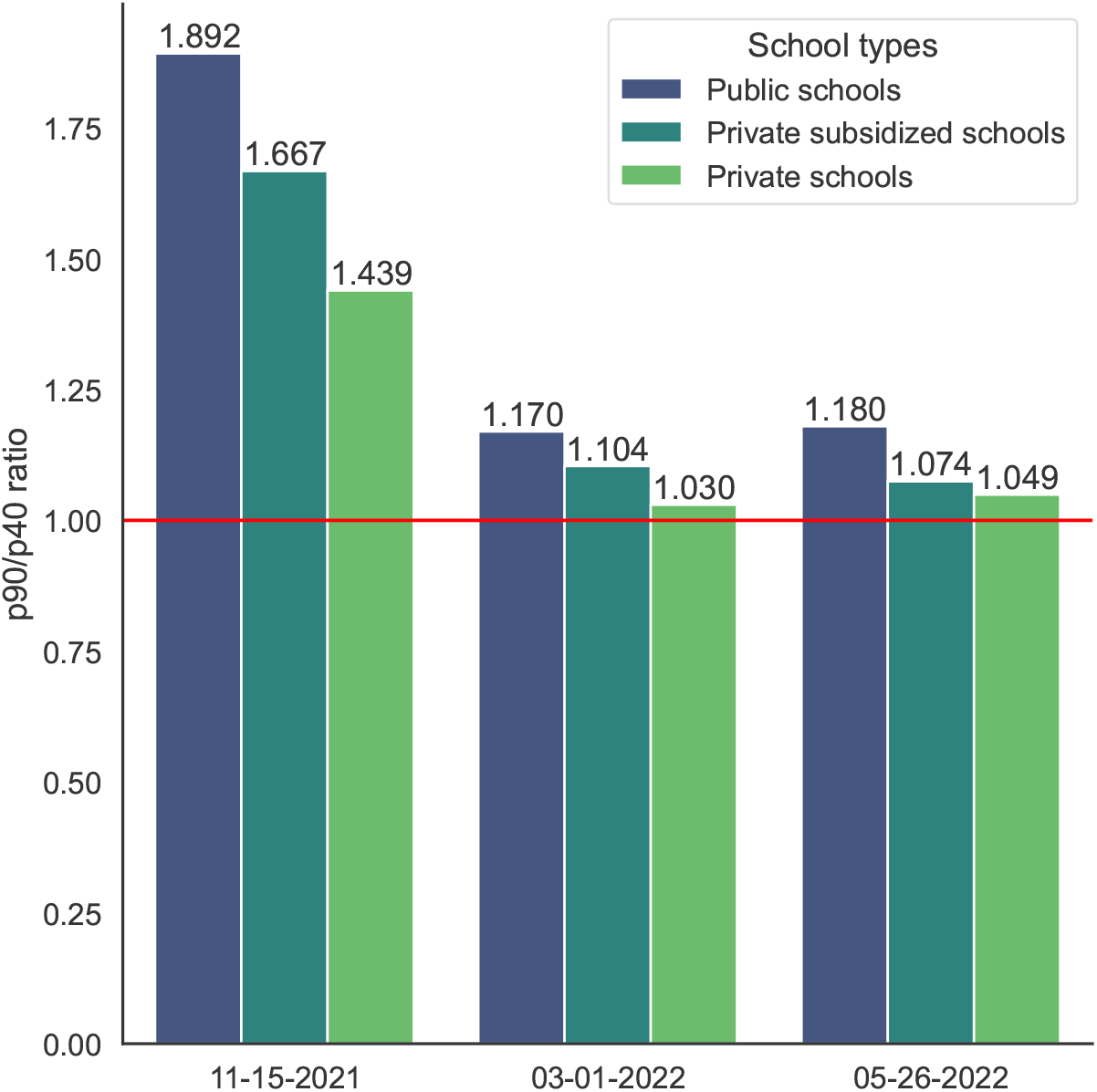
Inequity assessment of COVID-19 vaccination coverage in Santiago, Chile. The p90/p40 ratio is used to evaluate the inequity of the vaccination coverage for each school type at three time points along the COVID-19 pandemic: on November 15, 2021; March 1st, 2022; and May 26, 2022, covering the spread of different SARS-CoV-2 variants: *Delta, Omicron* and *Omicron BA*.*4-BA*.*5*, respectively. While inequity in vaccination coverage between schools types tend to diminish over time, it remains higher in public schools than that of the other types.

## 4 Discussion

This study evaluated the correlation between 9 SES indices and the COVID-19 vaccination coverage in schools belonging to 34 municipalities of the MA of Santiago, Chile; one of the countries with the highest rate of vaccination in the world.^27^ Subsequently to the evaluation of the correlation value between SES indices and the COVID-19 vaccination coverage using the Spearman’s correlation rank, we found that lower vaccination coverage is correlated with lower SES. Additionally, despite the fact that inequity in vaccination coverage diminished over time along the pandemic, this inequity remains highest in public schools compared to that of private and state-subsidized schools, for all municipalities (Figure 3). In general terms, our results are consistent with previous literature, indicating that vaccine coverage is lower in vulnerable populations living in poverty, having lower literacy and with a lesser educational level.^13, 26, 40^

Despite the fact that our results show a statistically significant correlation between the vulnerability of different municipalities and the COVID-19 vaccination coverage–i.e. municipalities with lower SES exhibit lower coverage–, different school types exhibit distinct vaccination coverage. Thus, both public and state-subsidized schools show lower vaccination coverage than that of private schools. Although the strength of these correlations varies depending on the municipality (Figure 2), a similar trend can be found along the pandemic (Figure 3): an important difference in vaccination coverage between school types, remains. On top of that, while analyzing the vaccination coverage in terms of percentages, as published by the Chilean government, reveal important differences between school types, examining the raw number of non-vaccinated children reveals an even worse and worrisome scenario: the number of children attending the schools with lower vaccination coverage. Importantly, the number of children assisting to public and state-subsidized schools surpass dramatically–more than five times–the number of children attending to private schools (873,354 vs 169,898, respectively). Moreover, the two groups with lower vaccination coverage represent up to 84% of the enrolled students in the MA of Santiago, Chile. Considering that municipalities with lower SES are the ones having the highest population density,^41^ a lower vaccination coverage together with higher population density, are both key factors to consider when dealing with the higher burden of COVID-19 on these populations.

One of the limitations of our study is that the SES indexes correspond to the municipal averages, so we cannot perform analyses nor extract conclusions at a granular level. In other words, assuming that an averaging SES index of the municipality would be an adequate descriptor for the whole spectrum of wealth exhibited on the municipality could be considered as a naive approach. This limitation becomes evident when trying to explain the differences in vaccination coverage between school types; we can only assume that the sub-populations that are enrolled in private schools (16%) are those that can afford the high tuition costs corresponding to the least vulnerable families in each municipality. Conversely, students enrolled in state-subsidized or in public schools are considered most vulnerable to the burden of COVID-19 (84%). Anyhow, without further socioeconomic details at an individual student level, we can only speculate about the causal roots of this inequity in vaccination coverage. Considering that COVID-19 vaccination in Chile is free of charge and governmentally orchestrated, determining the causal roots behind the low willingness to get vaccinated is crucial to develop better public policies. Available literature points to a combination of factors that may produce this uneven vaccination coverage. Among others, the lack of proper information about the importance of vaccination, insufficient incentives for vaccination, religious beliefs, the negative view of pharmaceutical companies, and low trust in the government, may all have played a role in this uneven vaccination rate.^9–11, 13, 26, 42^ Additionally, other physically impeding factors such as the availability of doses, ease of access to vaccination centers (determined in turn by location, availability of transport, size of waiting queues and opening hours) may also have an important role in determining the observed results. A more detailed analysis of these factors could be beneficial for designing improved vaccination campaigns and better public policies. However, if decisions continue to be made with coarse grained data, the syndemic character of COVID-19 and that of other future pandemics will be perpetuated.

## 5 Conclusion

In this study, we assessed the relationship between school vaccination coverage against COVID-19 and different socioeconomic variables defining the vulnerability of municipalities in the MA of Santiago, Chile. We show a strong correlation between several socioeconomic indices and vaccination coverage, confirming that the most vulnerable municipalities have lower vaccination coverage. We also show that these correlations were present in both public and state-subsidized schools. Unexpectedly, private schools have higher vaccination coverage exhibiting higher equity irrespective of the municipality SES. Furthermore, highest inequities in vaccination coverage reside in public education. Although this inequity decreased over time, it remains higher than that of other school types, namely, private and state-subsidized. Although we could not assume nor identify any causal root behind the lower vaccination coverage in vulnerable municipalities, available literature likely indicates that a combination of factors may be crucial to explain this behavior. Thus, the lack of proper information about the importance of vaccinations, insufficient incentives for children’s vaccination and low trust in the government may have all contributed to these low vaccination ratios. Even though we are unable to identify the causes of lower vaccination ratios among school-aged children in municipalities of lower SES in the MA of Santiago, Chile, it is imperative that public policies counteract this behavior. Otherwise, the inequity in vaccination coverage will perpetuate the harmful effects of the COVID-19 pandemic in the lower income population, exacerbating its syndemic nature.

## Data Availability

All data produced are available online at https://github.com/DLab/SantiagoSchoolVax

## 6 Acknowledgments

This material is based upon work supported by the U.S. Air Force Office of Scientific Research under award number FA9550-20-1-0196. Financial support is also acknowledged to Centro Ciencia & Vida, FB210008, Financiamiento Basal para Centros Científicos y Tecnológicos de Excelencia de ANID.

