## Supplemental Figures for "Lower vaccination coverage against COVID-19 in school-aged children is associated with low socioeconomic status in the Metropolitan Area of Santiago, Chile"

March 27, 2023

[†]These authors share first authorship
[*]Corresponding author

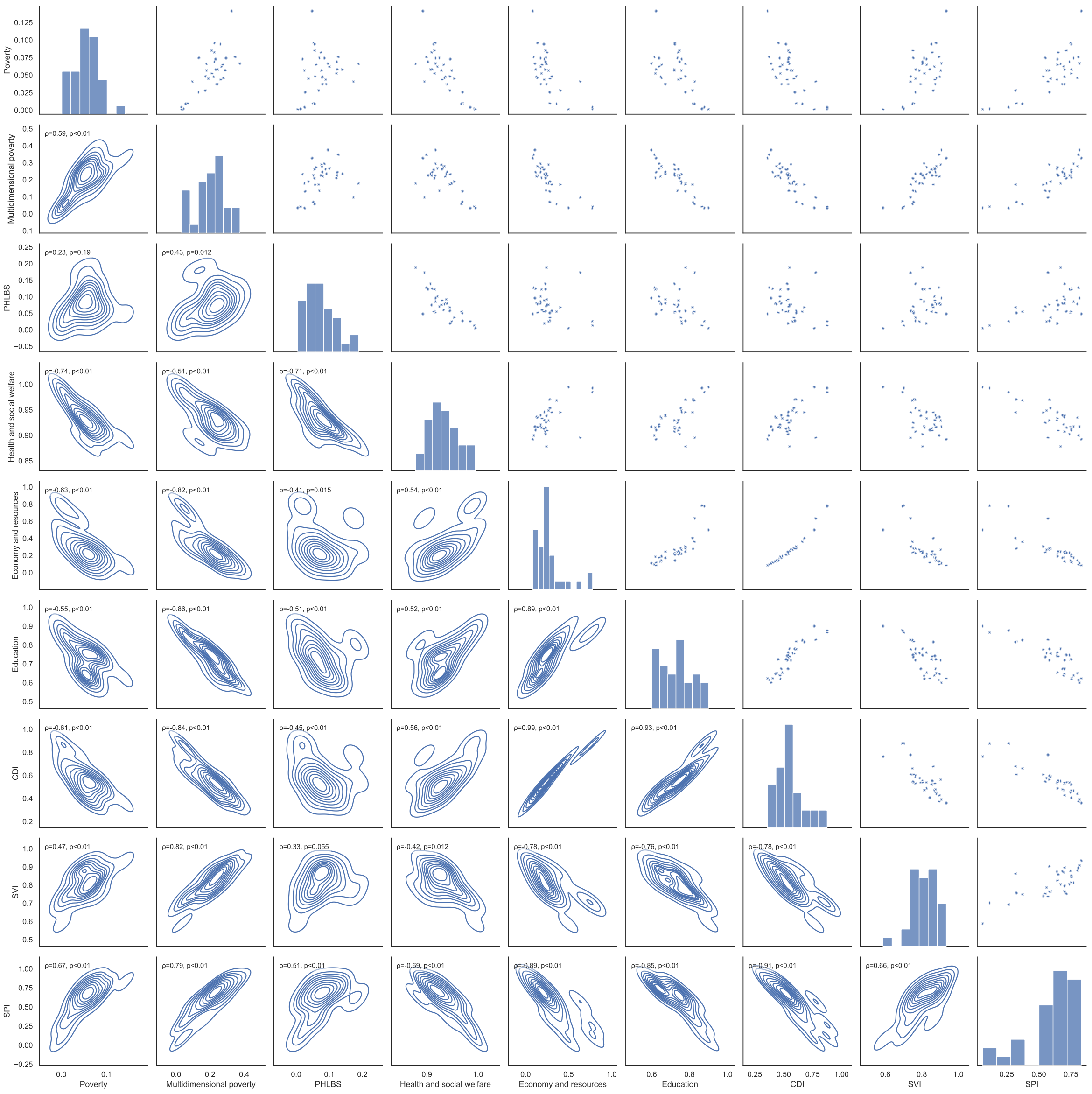


Figure S1: Correlation between different socioeconomic status (SES) indices in the Metropolitan Area of Santiago, Chile. A correlation between all SES indices is shown coupled with Spearman's rank correlation coefficient and its p-value.

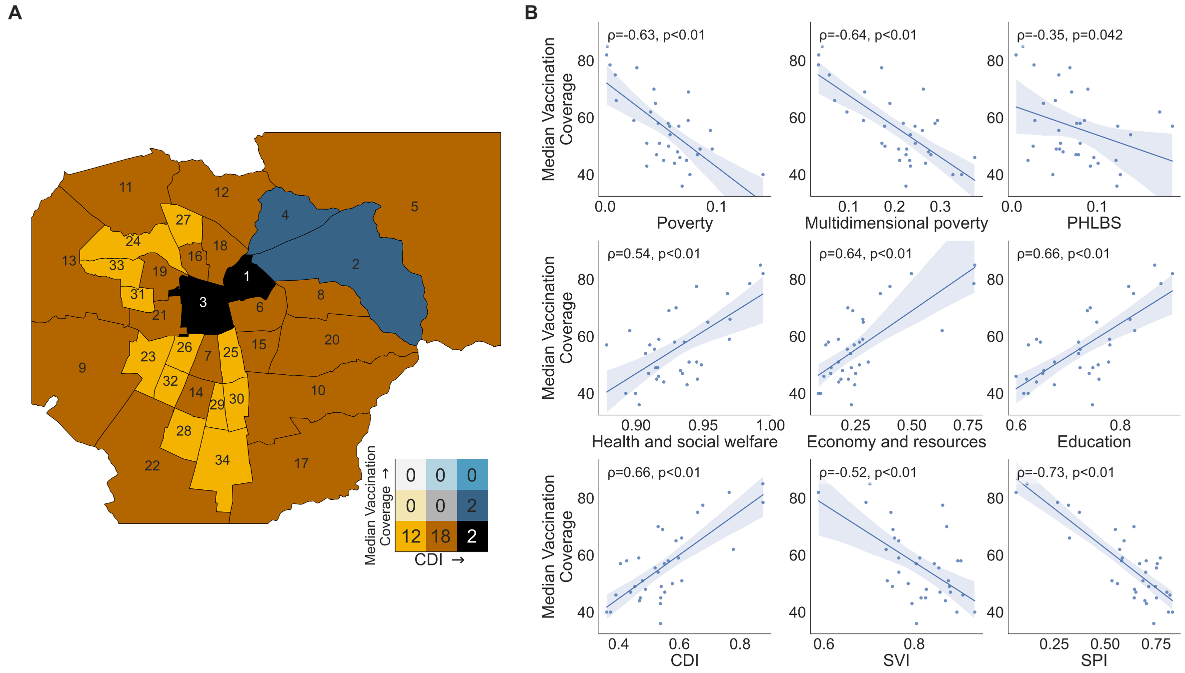


Figure S2: Correlation between school vaccination coverage and SES indices in the MA of Santiago, Chile as of Nov 15, 2021. Panel A), municipalities are colored according to their school vaccination coverage and their Community Development Index (CDI) in a Bivariate Map. Municipalities are numbered according to their rank in CDI from higher to lower values. The color coding is shown adjacent to the map using a 3 by 3 matrix, where the CDI increases from left to right and the vaccination coverage increases from bottom to top. The values are grouped into three levels. The number labeled in each municipality is sorted by decreasing ranking according to the CDI, a follows: 1, Providencia; 2, Las Condes; 3, Santiago; 4, Vitacura; 5, Lo Barnechea; 6, Ñuñoa; 7, San Miguel; 8, La Reina; 9, Maipú; 10, La Florida; 11, Quilicura; 12, Huechuraba; 13, Pudahuel; 14, La Cisterna; 15, Macul; 16, Independencia; 17, Puente Alto; 18, Recoleta; 19, Quinta Normal; 20, Peñalolén; 21, Estación Central; 22, San Bernardo; 23, Cerrillos; 24, Renca; 25, San Joaquín; 26, Pedro Aguirre Cerda; 27, Conchalí; 28, El Bosque; 29, San Ramón; 30, La Granja; 31, Lo Prado; 32, Lo Espejo; 33, Cerro Navia; 34, La Pintana. (B) The correlation value between nine SES Indexes and the median school vaccination coverage was evaluated in the 34 municipalities of the Metropolitan Area of Santiago, Chile. $\rho$ = Spearman's rank correlation coefficient; p = p-value.

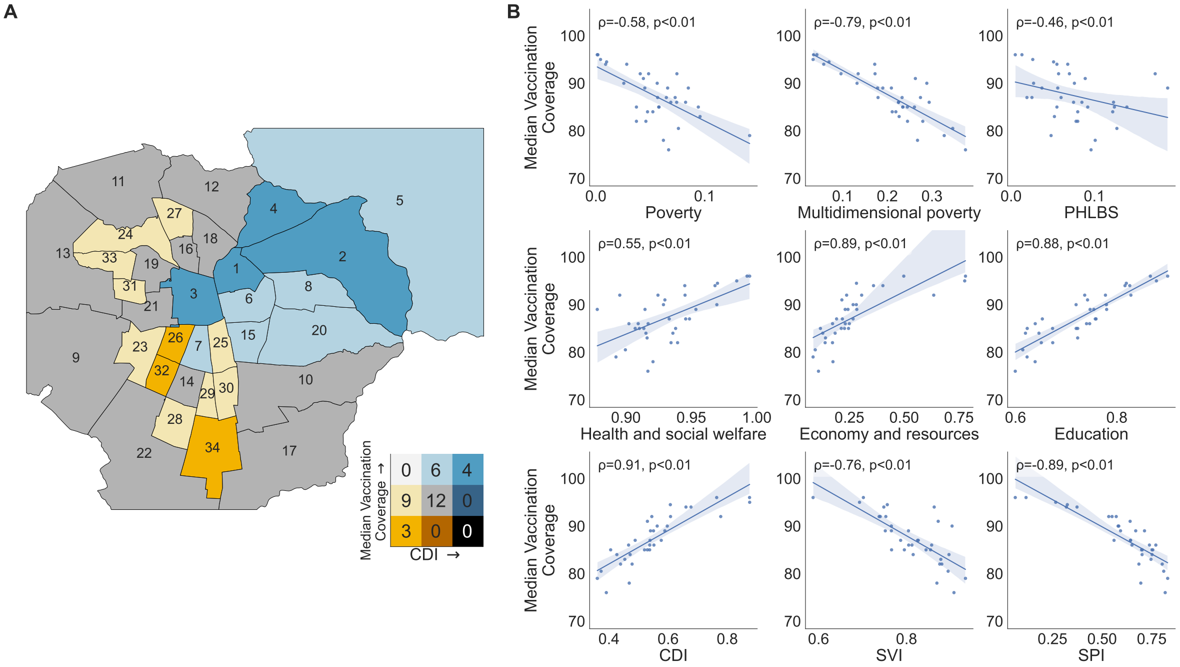


Figure S3: Correlation between school vaccination coverage and SES indices in the MA of Santiago, Chile as of Mar 1st, 2022. Panel A), municipalities are colored according to their school vaccination coverage and their Community Development Index (CDI) in a Bivariate Map. Municipalities are numbered according to their rank in CDI from higher to lower values. The color coding is shown adjacent to the map using a 3 by 3 matrix, where the CDI increases from left to right and the vaccination coverage increases from bottom to top. The values are grouped into three levels. The number labeled in each municipality is sorted by decreasing ranking according to the CDI, a follows: 1, Providencia; 2, Las Condes; 3, Santiago; 4, Vitacura; 5, Lo Barnechea; 6, Ñuñoa; 7, San Miguel; 8, La Reina; 9, Maipú; 10, La Florida; 11, Quilicura; 12, Huechuraba; 13, Pudahuel; 14, La Cisterna; 15, Macul; 16, Independencia; 17, Puente Alto; 18, Recoleta; 19, Quinta Normal; 20, Peñalolén; 21, Estación Central; 22, San Bernardo; 23, Cerrillos; 24, Renca; 25, San Joaquín; 26, Pedro Aguirre Cerda; 27, Conchalí; 28, El Bosque; 29, San Ramón; 30, La Granja; 31, Lo Prado; 32, Lo Espejo; 33, Cerro Navia; 34, La Pintana. (B) The correlation value between nine SES Indexes and the median school vaccination coverage was evaluated in the 34 municipalities of the Metropolitan Area of Santiago, Chile. $\rho$ = Spearman's rank correlation coefficient; p = p-value.

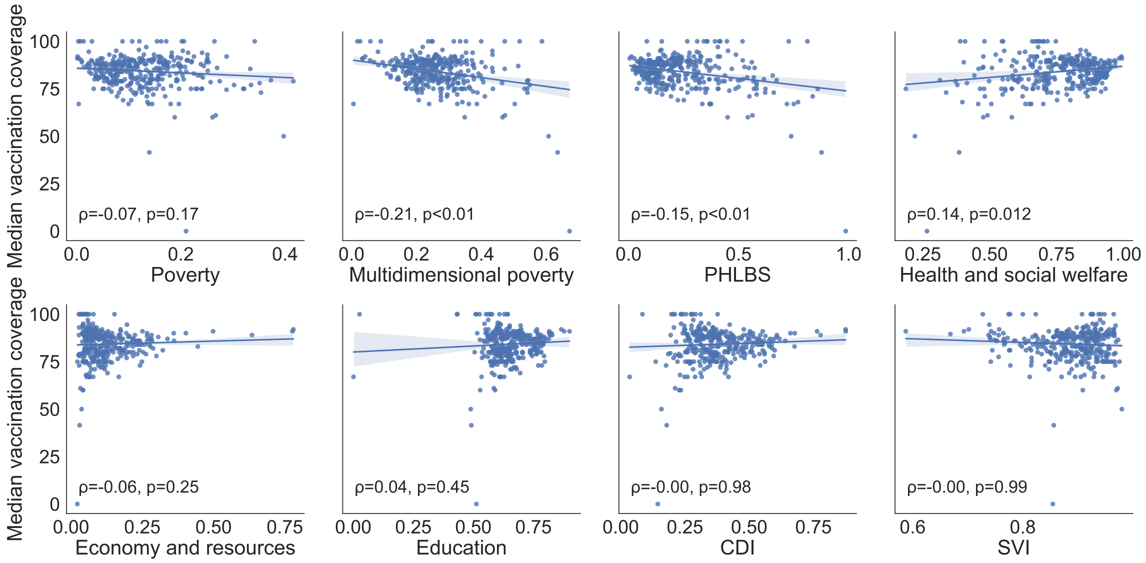


Figure S4: Correlation between school vaccination coverage and SES indices in Chile as of May 26, 2022. The correlation value between eight SES Indexes and the median school vaccination coverage was evaluated in the 345 municipalities of Chile. Each blue point represents one municipality. $\rho$ = Spearman's rank correlation coefficient; p = p-value. When considering the whole country, no correlations become evident. In contrast, when analyzing the MA of Santiago, Chile (see main text), several correlations between vaccination coverage and SES appear strongly, suggesting that municipalities in the MA of Santiago, Chile, are highly segregated by SES.
